# Sex differences in the associations of socioeconomic factors and cognitive performance with family history of Alzheimer’s disease

**DOI:** 10.1101/2024.06.12.24308850

**Authors:** Jun He, Brenda Cabrera-Mendoza, Eleni Friligkou, Adam P. Mecca, Christopher H. van Dyck, Gita A. Pathak, Renato Polimanti

## Abstract

**INTRODUCTION:** While higher socioeconomic factors (SEF) and cognitive performance (CP) have been associated with reduced Alzheimer’s disease (AD) risk, recent evidence highlighted that these factors may have opposite effects on family history of AD (FHAD).

**METHODS:** Leveraging data from the UK Biobank (N=448,100) and the All of Us Research Program (N=240,319), we applied generalized linear regression models, polygenic risk scoring (PRS), and one-sample Mendelian randomization (MR) to test the sex-specific SEF and CP associations with AD and FHAD.

**RESULTS:** Observational and genetically informed analyses highlighted that higher SEF and CP were associated with reduced AD and sibling-FHAD, while these factors were associated with increased parent-FHAD. We also observed that population minorities may present different patterns with respect to sibling-FHAD vs. parent-FHAD. Sex differences in FHAD associations were identified in ancestry-specific and SEF PRS and MR results.

**DISCUSSION:** This study contributes to understanding the sex-specific relationships linking SEF and CP to FHAD, highlighting the potential role of reporting, recall, and surviving-related dynamics.

## 1 BACKGROUND

With the increase of the aging population worldwide, Alzheimer’s disease (AD) has become one of the most challenging diseases of this century, because of its heavy medical and economic burden on individuals and society.^1^ In 2021, an estimated 56.9 million people suffered from AD and other dementias, indicating an age-standardized prevalence rate of 694.0 per 100,000 population.^2^ It is estimated that AD cases in preclinical, prodromal, and dementia stages comprised 416 million individuals, accounting for 22% of the total population aged 50 and older.^3^ Annually, 1.95 million deaths were attributed to AD and other dementias (25.2 per 100,000 population),^2^ which was the eighth leading cause of death worldwide.^4^ The estimated global economic burden of AD and related dementias has reached $2.8 trillion in 2019^5^. This figure was predicted to increase to $16.9 trillion in 2050,^5^ with the prevalence number expected to triple.^1^

AD is a complex disease attributable to biological and environmental influences, including genetic, psychosocial, cardiometabolic, and lifestyle factors.^6–18^ Among these, positive socioeconomic factors (SEF; e.g., higher income level and educational attainment) are demonstrated to be protective against AD.^6,9,11,16,17^ As a prodromal symptom, cognitive decline is a good predictor of clinical AD onset among older adults, independent of education and intelligence.^19–22^ However, in research of late-onset diseases such as AD, one difficulty is that even prospective studies with large sample sizes may not observe enough clinically diagnosed cases for robust analysis, due to the low incidence in middle-aged population.^23^ Therefore, researchers used family history of AD (FHAD) as a proxy of AD to improve statistical power, for instance in genome-wide association studies (GWAS).^24–26^ Several analyses reported that FHAD is associated with reduced cognitive performance (CP).^27–31^ However, a genetically informed study recently highlighted that investigating FHAD as a proxy for AD can introduce biases due to uncorrected survival bias and non-random participation in parental illness surveys.^32^ This raises important issues related to the dynamics linking SEF and CP to FHAD. Additionally, because of the known sex differences in AD risk^33^ and social determinants of health,^34^ it is crucial to assess whether these relationships may be influenced by sex-specific effects, differentiating FHAD assessment in females and males.

In the present study, we investigated sex differences in SEF and CP associations with AD and FHAD in two independent cohorts comprising a total of 688,419 participants. Specifically, we conducted observational and genetically informed analyses to explore how SEF (e.g., household income and educational attainment) and CP (e.g., fluid intelligence) are differentially associated with AD, sibling-FHAD, and parent-FHAD considering probands and parents’ sex.

## 2 METHODS

### 2.1 Study design

We investigated the associations of SEF and CP with FHAD by leveraging multiple analytical approaches (Figure 1). Firstly, we conducted an observational analysis using generalized linear regression models to estimate sex-specific SEF, CP, and *APOE*4 allele associations with AD, sibling-FHAD, mother-FHAD, and father-FHAD. To assess the dynamics related to parent-FHAD, we also estimated SEF, CP, and *APOE*4 allele associations after controlling for parental death and parental age at death. Using genetic data, we further investigated the associations of mother- and father-FHAD by performing polygenic risk score (PRS) and bidirectional one-sample Mendelian randomization (MR) analyses. In addition to investigating SEF and CP, our PRS analysis also tested the association of mother- and father-FHAD with brain imaging-derived phenotypes (IDPs). This permitted us to explore possible associations of parent-FHAD with brain structure and function.

**FIGURE 1.**
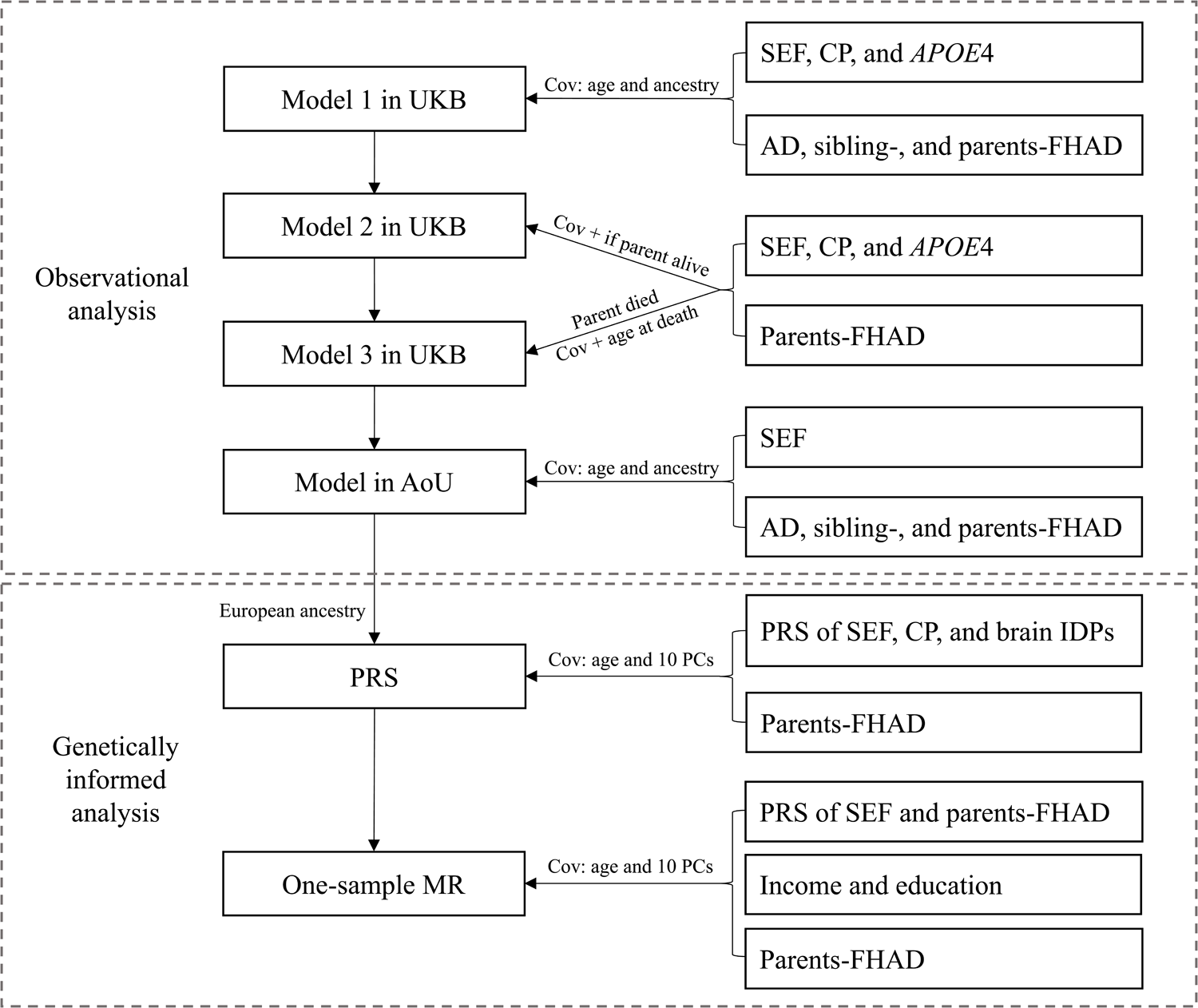
Flowchart of the analyses performed in the present study. UKB: the UK Biobank; AoU: the All of Us Research Program; Cov: covariates; SEF: socioeconomic factors; CP: cognitive performance; AD: Alzheimer’s disease; FHAD: family history of AD; PRS: polygenic risk score; PC: principal component; IDP: imaging-derived phenotype; MR: Mendelian randomization.

### 2.2 Data source

Phenotypic and genetic data were obtained from the UK Biobank (UKB)^35^ and the All of Us Research Program (AoU).^36^ UKB is a large population-based cohort including information regarding health conditions and genetic variation from participants recruited in the United Kingdom.^35^ AoU is a prospective, nationwide cohort study aiming to recruit more than one million participants in the United States.^36^ The sample investigated in the present study (Table 1) included 448,100 UKB participants (54% females; mean age at recruitment 56.7 years; median year of birth 1950) and 240,319 AoU participants (AoU Controlled Tier Dataset of Curated Data Repository version 7;^37^ 61% females, mean age at recruitment 54.5 years).

**TABLE 1.** Characteristics of female and male participants available from the UK Biobank and the All of Us Research Program.

### 2.3 Alzheimer’s disease and family history of Alzheimer’s disease

In UKB, AD was assessed with “first occurrence” information derived by combining primary care, hospital admission, death register, and self-reported data. Individuals with a first occurrence date of the ICD10 (Tenth Revision of the International Statistical Classification of Diseases and Related Health Problems) G30 (UKB Field ID: 131036) and F00 (UKB Field ID: 130836) codes were considered as AD cases. Sibling-, mother-, and father-FHAD were assessed through the UKB touchscreen questionnaire considering the “Alzheimer’s disease/dementia” reply to the following items, “Have any of your brothers or sisters suffered from any of the following illnesses?” (UKB Field ID: 20111), “Has/did your mother ever suffer from?” (UKB Field ID: 20110), and “Has/did your father ever suffer from?” (UKB Field ID: 20107).

In AoU, AD (SNOMED concept ID: 378419) was identified from the domain “conditions” using EHR data standardized by the Observational Medical Outcomes Partnership Common Data Model (OMOP CDM).^38^ FHAD information was collected using 2-level self-reported survey questions. In the first question, participants were asked: “Have you or anyone in your family ever been diagnosed with the following brain and nervous system conditions? Think only of the people you are related to by blood. Select all that apply.” (Participant-provided information, PPI concept ID: 43529272). If “Dementia (includes Alzheimer’s, vascular, etc.)” was selected from the optional diseases, then the second question was applied: “Including yourself, who in your family has had dementia (includes Alzheimer’s, vascular, etc.)? Select all that apply.” (PPI concept ID: 836807). Participants’ choices of “sibling”, “mother”, and “father” were used to define sibling-, mother-, and father-FHAD, respectively.

### 2.4 Socioeconomic factors and cognitive performance

In UKB, SEFs were assessed through self-reported information related to material deprivation, household income, and educational attainment. Specifically, material deprivation was assessed using the Townsend deprivation index (UKB Field ID: 189), which combines information regarding unemployment, non-car ownership, non-home ownership, and household overcrowding for any geographical area.^39^ This was calculated for each UKB participant based on the preceding national census output areas in which their postcode was located. Average total household income before tax (UKB Field ID: 738) was assessed in the touchscreen questionnaire item “What is the average total income before tax received by your HOUSEHOLD?”. This question provided five answer options: less than £18,000, £18,000 to £30,999, £31,000 to £51,999, £52,000 to £100,000, and greater than £100,000. UKB educational attainment was derived from three touchscreen questionnaire items: “At what age did you complete your continuous full time education?” (UKB Field ID: 845), “In which year did you first finish full-time education (school, college or university)?” (UKB Field ID: 22501), and “Which of the following qualifications do you have?” (UKB Field ID: 6138). With respect to CP, we leveraged information available from the UKB cognitive function assessment considering three items: mean time to correctly identify matching cards (UKB Field ID: 20023), maximum digits remembered correctly (UKB Field ID: 4282), and fluid intelligence score (UKB Field ID: 20016; a sum of the number of correct answers to 13 fluid intelligence questions). A description of the questions and tasks used in the UKB assessment is available at https://biobank.ndph.ox.ac.uk/showcase/.

In AoU, household income (PPI concept ID: 1585375) was determined through the survey question “What is your annual household income from all sources?” with nine answer options: “Less than $10,000”, “$10,000-$24,999”, “$25,000-$34,999”, “$35,000-$49,999”, “$50,000-$74,999”, “$75,000-$99,999”, “$100,000-$149,999”, “$150,000-$199,999”, and “$200,000 or more”. Educational attainment (PPI concept ID: 1585940) was assessed via the survey question “What is the highest grade or year of school you completed?” with eight options: “Never attended school or only attended kindergarten”, “Grades 1 through 4 (Primary)”, “Grades 5 through 8 (Middle school)”, “Grades 9 through 11 (Some high school)”, “Grade 12 or GED (High school graduate)”, “1 to 3 years after high school (Some college, Associate’s degree, or technical school)”, “College 4 years or more (College graduate)”, and “Advanced degree (Master’s, Doctorate, etc.)”. No cognitive assessment is currently available in AoU.

### 2.5 Genetic data

We leveraged individual-level genetic data available from UKB and AoU. UKB genetic information has been described previously.^35^ Briefly, UKB participants were genotyped using a custom Axiom array and imputed using the Haplotype Reference Consortium reference panel.^35^ In our analysis, we used quality control criteria and ancestry assignments performed by the Pan-UKB initiative.^40^ AoU genetic information was extracted from whole-genome sequencing data using the ACAF threshold callset.^41^ Quality control criteria and ancestry inference have been described elsewhere.^42^ Based on the dataset provided by AoU, we also removed variants with minor allele frequency (MAF) < 1%, call rate < 0.95, and Hardy-Weinberg equilibrium *P* < 1×10^-6^.

*APOE* genotypes were defined considering rs429358 and rs7412 single nucleotide polymorphisms (SNPs) as previously described.^43^ In our analysis, individuals were stratified into *APOE*4 carriers and non-carriers based on their genotypes.

To further investigate FHAD, SEF, and CP relationships, we also used sex-stratified genome-wide association statistics previously calculated using UKB cohort. Details regarding the quality control and the association analysis are available at https://github.com/Nealelab/UK_Biobank_GWAS. Briefly, the genome-wide association analysis was conducted using regression models available in Hail (available at https://hail.is/) and including the top-20 within-ancestry principal components (PC), age, and age^2^ as covariates. Because of the limited sample size available for other ancestry groups, we used genome-wide data generated from the analysis of 361,194 unrelated individuals of European descent. To explore FHAD associations with brain structure and function, we leveraged sex-stratified genome-wide association statistics of brain IDPs assessed in up to 33,224 individuals of European descent (52% females).^44^ Brain IDPs were derived from six MRI modalities: T1-weighted structural image, T2-weighted fluid-attenuated inversion recovery (FLAIR) structural image, diffusion MRI (dMRI), resting-state functional MRI (rfMRI), task functional MRI (tfMRI), and susceptibility-weighted imaging (SWI). Detailed information regarding imaging processing and quality control information is available at https://biobank.ctsu.ox.ac.uk/crystal/crystal/docs/brain_mri.pdf.

### 2.6 Observational analysis

We applied logistic regression models to UKB and AoU phenotypic data to investigate SEF and CP associations with AD and FHAD (sibling, father, and mother) in females and males. In UKB, three models were considered. In model 1, we estimated sex-specific associations of SEF, CP, and *APOE*4 status with AD and FHAD, adjusting for age and genetically inferred ancestry. In model 2, we assessed the same sex-specific relationships with father- and mother-FHAD, adjusting for age, genetically inferred ancestry, and parental death (father’s death (UKB Field ID: 1797) for father-FHAD and mother’s death (UKB Field ID: 1835) for mother-FHAD). In model 3, the sex-specific association analysis was restricted to participants whose parent died (participants whose father died in father-FHAD analysis and participants whose mother died in mother-FHAD analysis) and included age, genetically inferred ancestry, and parental age at death (father’s age at death (UKB Field ID: 1807) for father-FHAD and mother’s age at death (UKB Field ID: 3526) for mother-FHAD). Bonferroni correction was applied to account for the number of tests performed (*P <* 8.33×10^-4^ in model 1 and *P <* 1.67×10^-3^ in model 2 and 3). Variables surviving multiple testing correction were then entered into a multivariable model to estimate whether their sex-specific effects were independent of each other. Additionally, we estimated sex differences by comparing sex-specific association statistics using z-test.

In AoU, we tested the sex-specific association of household income and educational attainment with AD and sibling-, father-, and mother-FHAD including age and genetically inferred ancestry as covariates. Because no information regarding parental death and parental age at death is available in AoU, we could not consider other models. However, because AoU is an ancestrally diverse cohort, we conducted an ancestry stratified analysis to examine whether SEF association with AD and FHAD relationships were consistent across populations. In both cross-ancestry and ancestry-stratified analyses, Bonferroni correction was applied to account for the number of tests performed. All variables were subsequently entered into a multivariable regression model to assess their independent effects.

### 2.7 Polygenic risk scoring

PRS analysis was employed to investigate SEF, CP, and brain-IDP sex-specific associations with parent-FHAD. Specifically, we derived PRS from previously calculated UKB genome-wide association statistics (see 2.5 Genetic data) and then tested them with respect to parent-FHAD in AoU. As mentioned above, this analysis was limited to European-descent individuals, because of the limited sample size available for other population groups. PRS were estimated using Polygenic Risk Score–Continuous Shrinkage (PRS-CS)^45^ and UKB European-ancestry participants as the LD reference panel^46^. Then, PRS-CS weights were used to estimate individual PRS for AoU participants of European descent via PLINK 1.9 --score command.^47^ PRS associations were estimated via logistic regression with the PRS as the independent variable, mother- or father-FHAD as the dependent variable, and age and the top 10 within-ancestry PCs as covariates. With respect to SEF and CP, we tested PRS related to Townsend deprivation index, household income, age completed full time education, education qualification, year ended full time education, mean time to correctly identify matches, maximum digits remembered correctly, and fluid intelligence score. To account for the number of PRS tests performed, we applied a Bonferroni correction (*P* < 1.56×10^-3^).

In addition to SEF and CP PRS, we also performed an exploratory analysis testing sex-specific brain-imaging PRS (see 2.5 Genetic data) with respect to mother- and father-FHAD. The PRS were derived only for brain IDP GWAS with SNP-based heritability Z > 7 in both females and males (Supplementary Table S1). Due to the limited sample size of IDP GWAS (up to 33,224), we considered this as an exploratory analysis and used a relaxed significance threshold (*P* < 0.01) for both univariable and multivariable analyses. SEF and CP PRS surviving multiple testing and nominally significant brain-imaging PRS were then entered in a multivariable regression model to assess their independent effects.

### 2.8 One-sample Mendelian randomization

To investigate further the sex-specific dynamics linking SEF (i.e., educational attainment and household income) to parent-FHAD, we performed a bidirectional one-sample MR analysis in females and males separately^48,49^. Using PRS as instrumental variables, we performed an MR analysis in AoU European-ancestry participants by applying the two-stage predictor substitution (TSPS) estimator available in *OneSampleMR* package in R.^50^ Age and the top 10 within-ancestry PCs were included as covariates. When the outcome in the model was a binary variable, logistic regression was used in the second stage; when the outcome was a quantitative variable, linear regression was used. Bonferroni correction (*P* < 0.005) was applied to account for the number of MR tests performed. To ensure the reliability of our findings, we also recalculated MR estimates using the two-stage least-squares (2SLS) estimator available in *ivreg* package in R. Finally, we conducted weak-instrument and Wu-Hausman tests to examine whether the instrumental PRS was weakly correlated with the exposure variables and whether the 2SLS estimator was consistent with the ordinary least squares (OLS) estimator.

## 3 RESULTS

### 3.1 Observational analysis

After adjusting for age and genetically inferred ancestry, high SEF and CP were associated with reduced AD and sibling-FHAD, while their relationship with father- and mother-FHAD was reversed (i.e., high SEF and CP were associated with increased parent-FHAD; Figure 2, Supplementary Table S2). For instance, with respect to SEF, high household income was associated with reduced AD (β_female_ = −0.236, *P* = 1.54×10^-15^; β_male_ = −0.267, *P* = 3.05×10^-24^) and sibling-FHAD (β_female_ = −0.148, *P* = 7.20×10^-7^; β_male_ = −0.089, *P* = 0.003) but increased mother-FHAD (β_female_ = 0.086, *P* = 2.98×10^-33^; β_male_= 0.077, *P* = 3.59×10^-24^) and father-FHAD (β_female_ = 0.092, *P* = 1.29×10^-22^; β_male_ = 0.096, *P* = 5.51×10^-22^). This dynamic was consistent also when considering negative SEF such as not having any education qualifications, which was associated with increased AD (β_female_ = 0.393, *P* = 4.05×10^-17^; β_male_ = 0.365, *P* = 1.57×10^-13^) and sibling-FHAD (β_female_= 0.406, *P* = 5.09×10^-13^; β_male_ = 0.426, *P* = 9.60×10^-11^), but reduced mother-FHAD (β_female_= −0.333, *P* = 9.10×10^-65^; β_male_ = −0.303, *P* = 8.08×10^-42^) and father-FHAD (β_female_ = −0.363, *P* = 2.65×10^-37^; β_male_ = −0.375, *P* = 1.84×10^-31^). CP measurements showed the same association pattern. For example, high fluid intelligence was associated with reduced AD (β_female_ = −0.169, *P* = 1.03×10^-12^; β_male_ = −0.153, *P* = 3.62×10^-12^) and sibling-FHAD (β_female_ = −0.047, *P* = 0.030; β_male_ = −0.070, *P* = 0.003) but increased mother-FHAD (β_female_ = 0.065, *P* = 9.09×10^-31^; β_male_ = 0.047, *P* = 1.31×10^-15^) and father-FHAD (β_female_ = 0.040, *P* = 6.59×10^-8^; β_male_ = 0.054, *P* = 2.31×10^-12^). Differently from SEF and CP, *APOE*4 carrier status showed increasing effects on AD, sibling-FHAD, mother-FHAD, and father-FHAD (Figure 2, Supplementary Table S2). When comparing female and male effects, we observed a sex difference only for *APOE*4 carrier status on AD (β_female_ = 1.676 *vs*. β_male_ = 1.264, sex-difference *P* = 4.95×10^-10^). When we entered Bonferroni significant variables from the univariable analyses into multivariable models, SEF and CP associations were strongly attenuated due to their overlap, but their effect directions were consistent with what was observed in the univariable analyses (Supplementary Table S3). To further investigate SEF and CP relationships with parent-FHAD, we tested two additional models. In Model 2, controlling for parental death (father’s death for father-FHAD and mother’s death for mother-FHAD) in addition to age and genetically inferred ancestry, we observed no substantial difference with respect to the Model-1 effects (Supplementary Figure S1, Supplementary Tables S4 and S5). In Model 3, restricting the analysis to participants whose parent died (father died in father-FHAD analysis and mother died in mother-FHAD analysis) and including age, genetically inferred ancestry, and parental age at death as covariates, we observed a reduction in the effect size observed with several results becoming null. However, for those surviving Bonferroni multiple testing correction, the effect direction was consistent with Model 1 (Supplementary Figure S1, Supplementary Tables S6 and S7).

**FIGURE 2.**
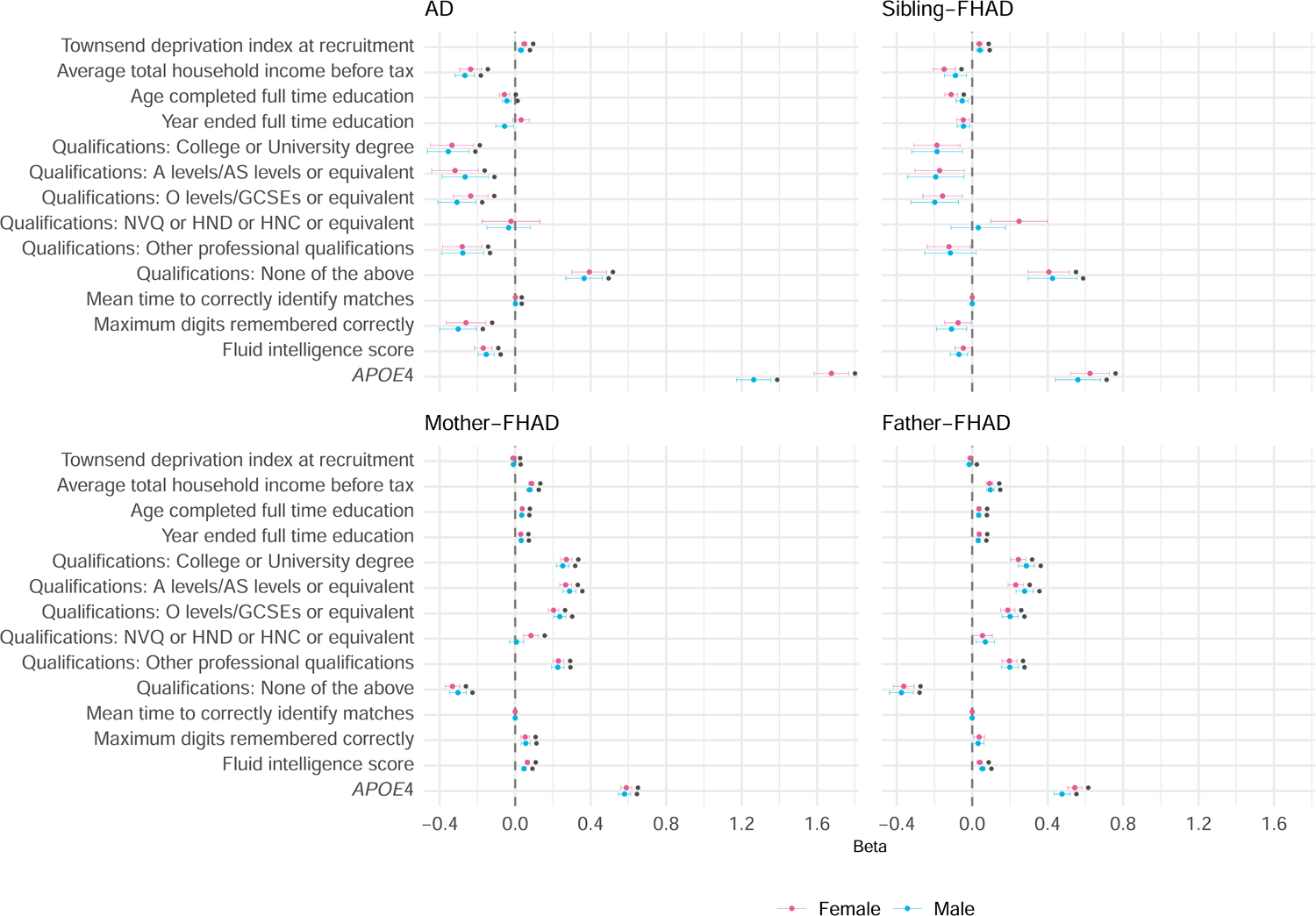
Associations of socioeconomic factors, cognitive performance, and *APOE*4 status with FHAD in the UK Biobank. Full results are available in Supplementary Table S2. Star symbols indicate associations surviving Bonferroni multiple testing correction. AD: Alzheimer’s disease; FHAD: family history of AD.

In the AoU cohort, high educational attainment and income were also associated with reduced AD and with increased parent-FHAD in both sexes (Supplementary Figure S2, Supplementary Tables S8 and S9). Null effects were observed with respect to sibling-FHAD (*P* > 0.05). Leveraging AoU population diversity, we also investigated ancestry-specific associations (Figure 3; Supplementary Table S10). Controlling for educational attainment and income and applying a Bonferroni correction accounting for the number of tests performed (*P <* 2.5×10^-3^), genetically inferred African descent (AFR) was associated with increased sibling-FHAD in females (β = 0.412, *P* = 0.002), while an inverse relationship was present with respect to father-FHAD in both females (β = −0.444, *P* = 8.2×10^-9^) and males (β = −0.539, *P* = 2.72×10^-4^). AFR descent was also inversely associated with mother-FHAD in males (β = −0.517, *P* = 8×10^-6^), but the effect was statistically smaller in females (β = −0.113, *P* = 0.043; sex-difference *P* = 0.002). In males, mother-FHAD was also inversely associated with genetically inferred Central/South Asian (CSA; β = −0.953, *P* = 3.19×10^-4^). Conversely, mother-FHAD was inversely associated with genetically inferred East Asian descent in females (EAS; β = −0.477, *P* = 8.02×10^-4^) but not in males (*P* = 0.627). We also conducted an ancestry-stratified analysis, testing income and educational attainment associations with AD and FHAD. In line with the results obtained in the whole AoU cohort, these factors were associated with reduced AD and increased parent-FHAD (Supplementary Tables S11 and S12).

**FIGURE 3.**
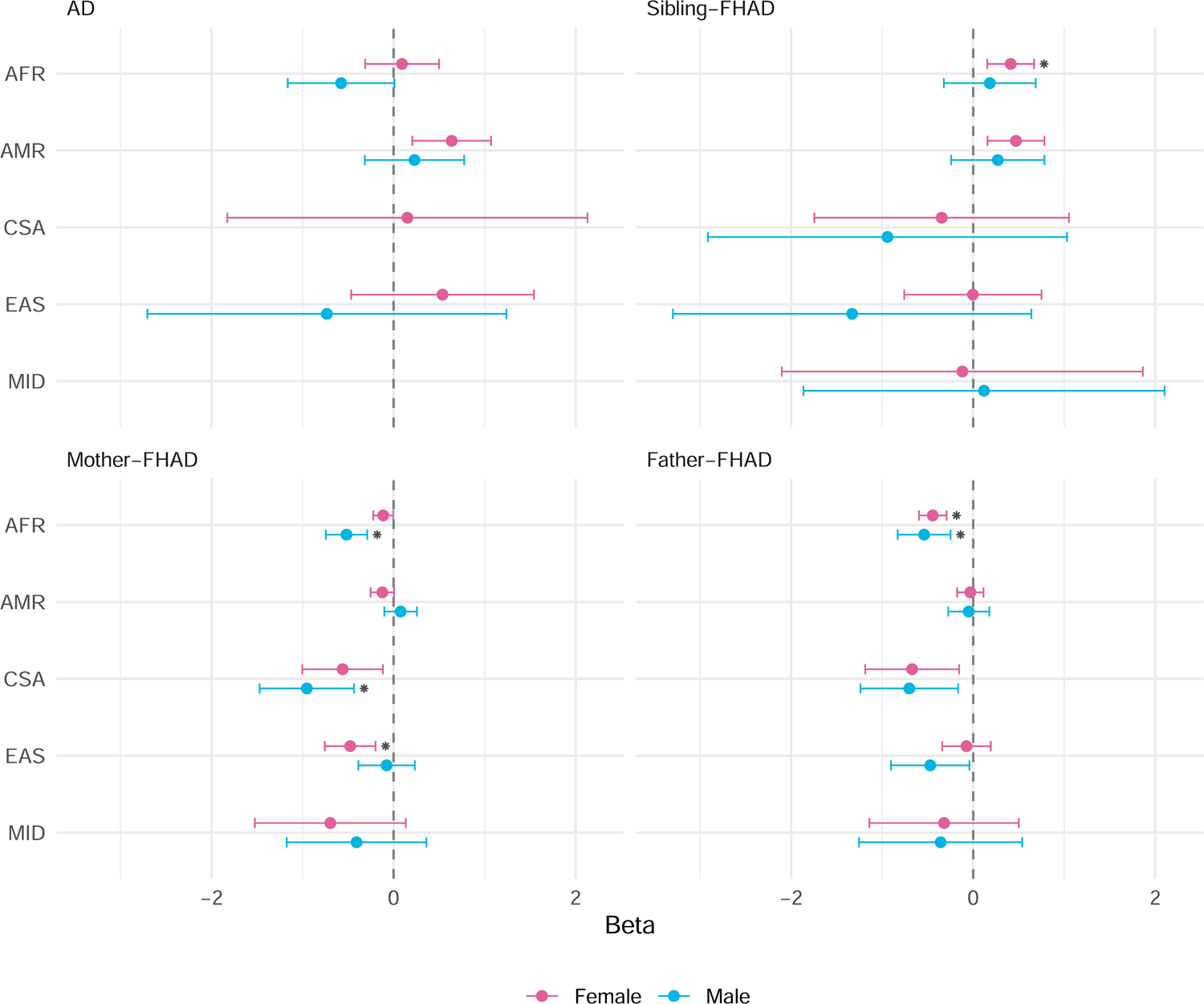
Associations between genetically inferred ancestry and FHAD in the All of Us Research Program. AD betas in CSA (males) and MID (both sexes) are not shown due to extremely small sample sizes. Full results are available in Supplementary Tables S10. AD: Alzheimer’s disease; FHAD: family history of AD; EUR: European ancestry; AFR: African ancestry; AMR: Admixed American descent; CSA: Central/South Asian ancestry; EAS: East Asian ancestry; MID: Middle Eastern ancestry.

### 3.2 Polygenic risk scoring

To further investigate parent-FHAD, we leveraged UKB GWAS to derive SEF, CP, and FHAD PRS and tested them in AoU cohort. In both males and females, the strongest parent-FHAD associations were related to PRS derived from “Illnesses of mother: Alzheimer’s disease/dementia” and “Illnesses of father: Alzheimer’s disease/dementia” (Figure 4, Supplementary Table S13). The direction of the association was in line with the expectation that high parent-AD PRS would be associated with increased parent-FHAD. Interestingly, in both sexes we observed cross-parent PRS associations with mother-FHAD PRS associated with father-FHAD and vice versa (Figure 4, Supplementary Table S13). However, there was a sex difference in the association of mother-AD PRS with father-FHAD, which was stronger in males (β = 2.028, *P* = 1.08×10^-11^) than in females (β = 1.004, *P* = 1.07×10^-5^; sex-difference *P* = 0.006). Considering Bonferroni-significant SEF PRS associations (*P* < 1.56×10^-3^), we observed the same pattern of the observational analyses described above, where PRS related to positive SEF were associated with increased parent-FHAD (Figure 4, Supplementary Table S13). For instance, having a college or university degree was associated with increased father-FHAD in both females (β = 0.687, *P* = 2.12×10^-6^) and males (β = 0.677, *P* = 3.69×10^-4^). While several SEF PRS were Bonferroni significant only in one sex but only nominally replicated or null in the other (Figure 4, Supplementary Table S13), we observed only one statistically significant sex difference. Townsend deprivation index PRS was associated with reduced mother-FHAD in males (β = −0.784, *P* = 0.001), but not in females (β = 0.034, *P* = 0.868; sex-difference *P* = 0.009). None of the CP PRS survived Bonferroni multiple testing correction, but the effect direction of the nominally significant results was consistent with the observational findings described above (Supplementary Table S13).

**FIGURE 4.**
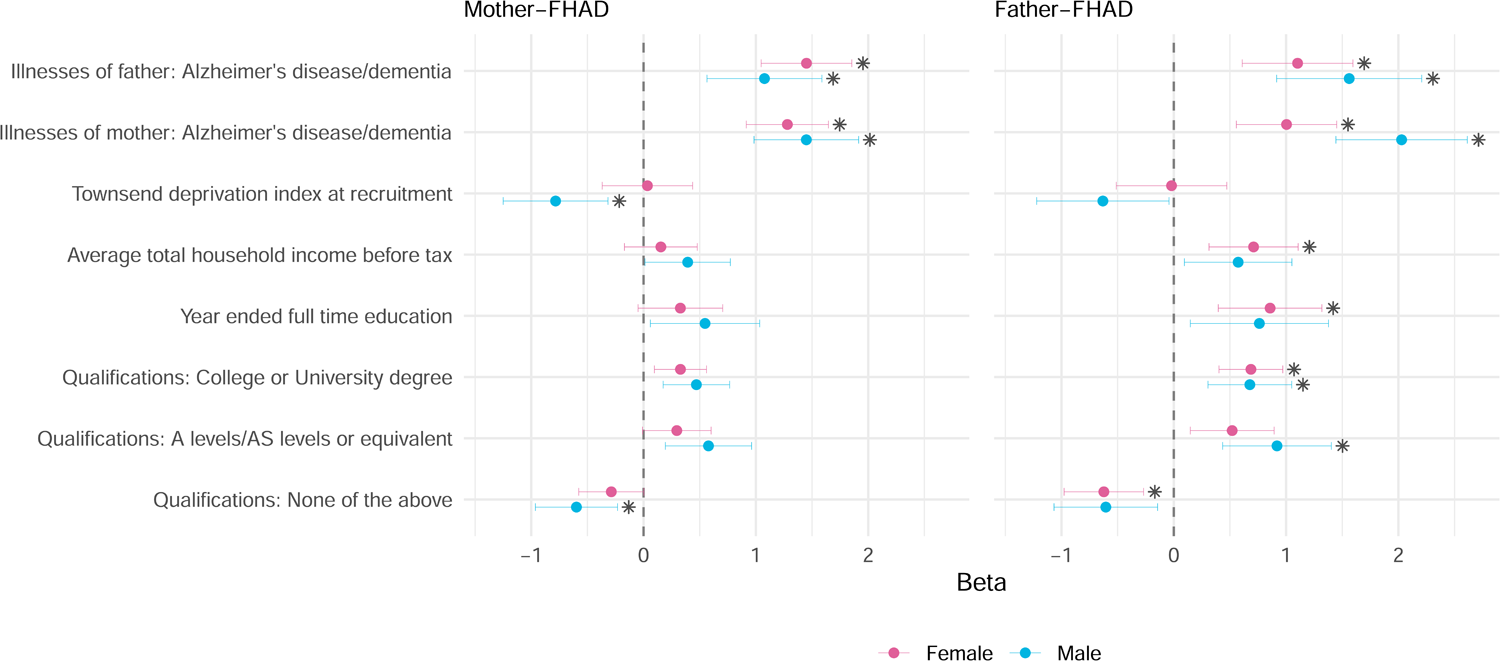
Associations of polygenic risk scores related to socioeconomic factors and cognitive performance with FHAD. Full results are available in Supplementary Table S13. Star symbols indicate associations surviving Bonferroni multiple testing correction. FHAD: family history of Alzheimer’s disease.

To explore FHAD relationship with brain structure and function, we investigated PRS derived from brain IDPs (Supplemental Table S14). Considering a relaxed significance threshold (*P* < 0.01), we observed that PRS related to the mean diffusion tensor mode in anterior limb of the left internal capsule on fractional anisotropy skeleton (IDP 1544, dMRI TBSS MO Anterior limb of internal capsule L) was associated with reduced father-FHAD in females (β = −0.160, *P* = 0.009), but not in males (β = 0.127, *P* = 0.103; sex-difference *P* = 0.004). This brain IDP also showed a nominally significant sex difference with respect to mother-FHAD, but with opposite effect directions (β_female_ = 0.099, *P* = 0.049; β_male_ = −0.08, *P* = 0.195; sex-difference *P* = 0.025). Finally, we conducted a multivariate analysis of SEF, FHAD, and brain IDP PRS, which confirmed the independence and the direction of the effects detected in the initial PRS analyses (Supplementary Table S15).

### 3.3 One-Sample Mendelian randomization

Using the TSPS approach to perform a one-sample MR analysis, we investigated the bidirectional relationship between SEF and parent-FHAD. In both sexes, we observed that genetic liability to mother-FHAD affected father-FHAD and vice versa (Figure 5, Supplementary Table S16). The results derived using the TSPS method were highly consistent with the analyses obtained from the 2SLS approach (Supplementary Table S17). With both methods, female genetic liability to father-FHAD affected mother-FHAD more strongly than female genetic liability to mother-FHAD affected father-FHAD (TSPS difference-*P* = 0.038, Supplementary Table S16; 2SLS difference-*P* = 0.008, Supplementary Table S17). We observed Bonferroni significant effects of educational attainment and household income on father-FHAD in females and of educational attainment on mother-FHAD in males (Figure 5; Supplementary Table 16). The sensitivity analyses confirmed the reliability of the one-sample MR results, highlighting the lack of weak instrument bias and indicating that the outcomes tested are associated with the variables in our models (Supplementary Table S17).

**FIGURE 5.**
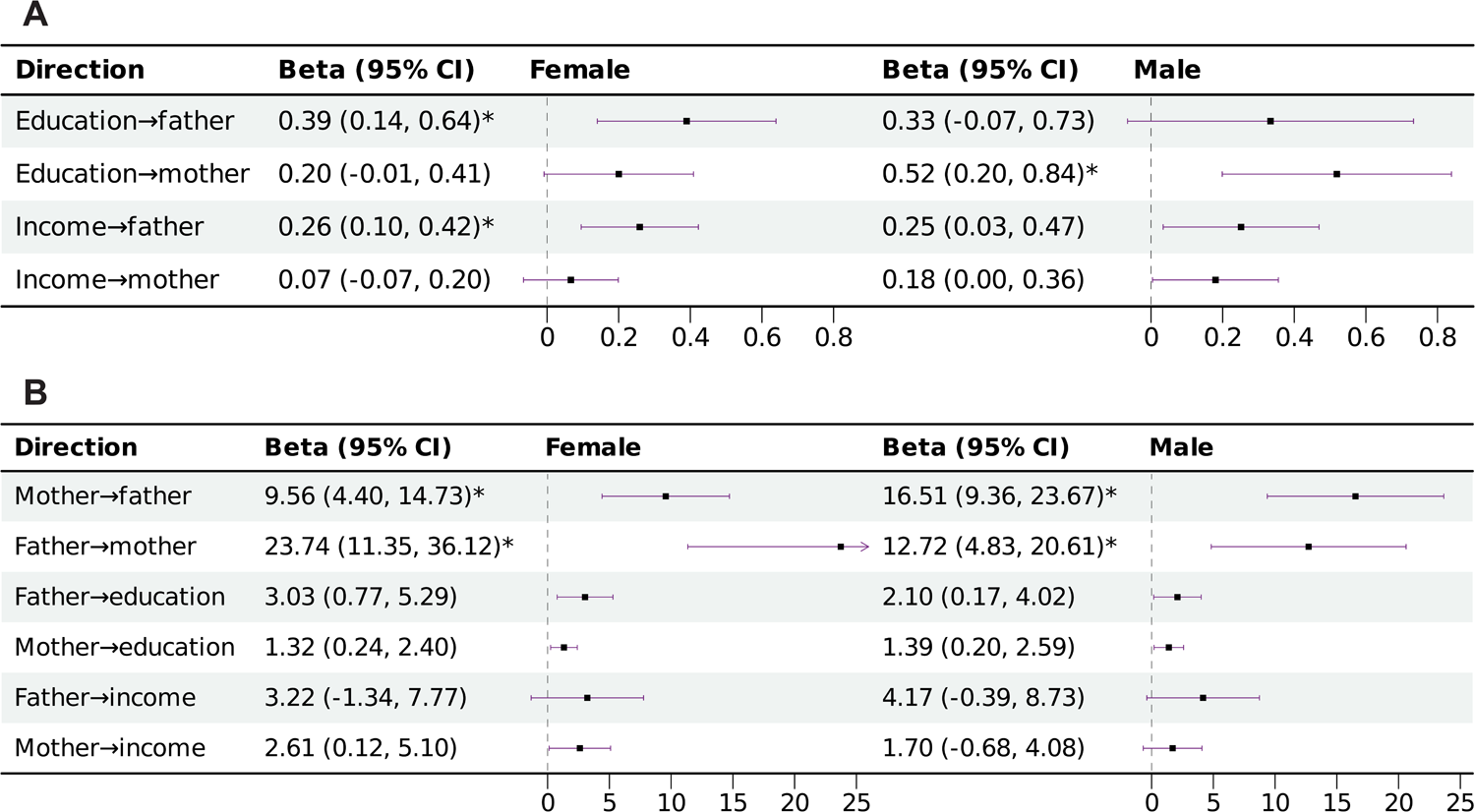
Bidirectional relationships between socioeconomic factors and family history of Alzheimer’s disease. (A) Causal effects of household income and educational attainment on parent-FHAD. (B) Causal effects of parent-FHAD. The two-stage predictor substitution estimator was used to assess the causal relationships. Full results are available in Supplementary Table S16. Star symbols indicate associations surviving Bonferroni multiple testing correction.

## 4 DISCUSSION

The prevalence of AD has been rising in the 21^st^ century due to increased life expectancy and an aging population, resulting in an enormous disease and economic burden.^5,51^ FHAD has been used in studies of AD to identify its genetic basis and associated factors with the aim of expanding sample sizes and improving statistical power.^23–26^ Recently, concerns have been raised about the accuracy of the self-reported parental information.^32^ In the present study, we used phenotypic and genetic data from two large cohorts to investigate sex-specific associations of SEF and CP with AD and FHAD, also exploring differences across population groups. In line with previous evidence,^6,11,22^ we found that high SEF and CP are associated with reduced AD. However, while the effect was consistent for sibling-FHAD, we observed an inverse relationship with respect to parent-FHAD: UKB and AoU participants with better SEF and/or CP reported increased father- and mother-FHAD compared to those with poor SEF and/or CP. There are several possible explanations for this unexpected outcome. One is reporting bias, which is prevalent in studies related to self-reported information and varies across different population groups, e.g., individuals of lower SEF and CP may be less likely to report accurate information.^52^ Regarding family history, the bias varies among family members; for example, the number of self-reported paternal family histories of cancer was reported to be significantly lower than that of maternal family histories.^53^ The second potential bias is misclassification.^54^ As material deprivation and SEF are important determinants of healthcare service accessibility and are intergenerationally transmissible, low-income individuals are more likely to have economical insurance or be uninsured.^55,56^ Consequently, the limited access to medical services and the low frequency of visits lead to underestimates of AD diagnosis of parents. Third, there may be recall bias: self-reported records could suffer more from recall bias in a population with cognitive decline.^57^ Therefore, those with poor CP may report a lower proportion of true FHAD.^58^ Fourth, survival bias may play an important role in these associations. It has been recognized that SEF is associated with all-cause mortality.^59^ In this way, parents of individuals in lower SEF would die early due to competing risks before they had a chance to develop AD.^60^ This is consistent with our finding that the associations substantially attenuated when we restricted the analysis to participants whose father or mother was deceased. Parental age could also be a confounder. Typically, the age at birth of individuals is correlated with socioeconomic status.^61^ Thus, those parents in families with lower SEF will be younger, resulting in a relatively lower prevalence rate of FHAD.

However, these dynamics may not affect sibling-FHAD, because they lived at the same period as participants and may have less survival bias and recall bias. Unlike the confounding bias of parental age, sibling’s age may not contribute to the bias substantially after adjusting for participants’ age. In the observational analysis in UKB cohort, *APOE*4 carrier status was consistently a risk factor for both FHAD and AD, as expected.^43^ Because this is a biological risk factor, it is less likely to be affected by the biases related to self-reported family history. However, we still observed that *APOE*4 effect on AD was larger than that on FHAD. With respect to biological factors, in our exploratory analysis regarding brain structure and function, we identified one IDP associated with FHAD in females. In line with previous AD studies,^62,63^ mean diffusion tensor mode in the left anterior limb of internal capsule (IDP dMRI TBSS MO Anterior limb of internal capsule L) was negatively associated with father-FHAD. However, the association of this brain IDP was not consistent between parents, as it was positively associated with mother-FHAD. This suggests that parent-specific effects may act on the biases discussed above.

Importantly, we found sex differences in AD and FHAD associations. In the UKB observational analysis, *APOE*4 was more strongly associated with AD in females than in males, consistent with the finding that females with this allele have a greater risk of developing AD than male *APOE*4 carriers.^64^ In AoU, we identified multiple sex differences when investigating FHAD associations among population groups. After controlling for educational attainment and income, AFR and CSA male participants were less likely to report mother-FHAD than females. This sex-specific association was reversed in EAS where females were less likely to report mother-FHAD than males. This suggests that the sex-specific interplay of the biases discussed above with disparities in healthcare access, life expectancy, and AD prevalence observed in population minorities.^65,66^

Sex differences were also present in the results of our genetically informed analyses. Specifically, mother-AD PRS was more strongly associated with father-FHAD in males than in females. Additionally, one-sample MR analysis highlighted that female genetic liability to father-FHAD affected mother-FHAD more strongly than female genetic liability to mother-FHAD affected father-FHAD. These differences could be related to parental differences in the perception of family history. For example, females reported more obligation to care for their mother than their father, and mother-daughter dyads engaged in more mutually open conversations with less conflict.^67^ Moreover, females might know more about their mother’s health than their father’s, and maternal family history was more strongly associated with AD risk factors than paternal family history.^68^ Another possible aspect contributing to the sex differences in FHAD genetics is assortative mating (mate choice driven by phenotypic similarity). Recently, assortative mating, SEF, and participation bias have been reported in the context of the polygenic risk of neuropsychiatric and behavioral traits.^69^ Based on this previous evidence and our current results, we hypothesize that the interplay between FHAD and SEF could contribute to sex-specific assortative mating.

Strengths of the present study include the use of phenotypic and genetic data from two large cohorts to investigate the sex-specific associations of SEF and CP with AD and FHAD. This permitted us to triangulate robust evidence integrating multiple approaches and data types. However, we need to acknowledge several limitations. First, because of the UKB and AoU questionnaires, we could not distinguish FHAD from family history of other dementias in our analysis. However, AD is the most common form of dementia, accounting for 60 to 80% of the cases,^70^ implying that our findings are mainly driven by FHAD. Although our analyses accounted for sex, age, genetically inferred ancestry, parental death, and parental age at death, we could not control for other factors. For example, we did not include psychosocial factors, lifestyle, and healthcare accessibility of the first-degree relatives, which may contribute to the biased associations found in this study. Finally, UKB and AoU are not cohorts with randomly selected participants and they are not representative of the general population, which may introduce volunteer bias as an additional source of confounding, thus affecting the estimated associations.^71^

In conclusion, the present study contributes to the understanding of how SEF and CP can affect parent FHAD differently than AD and sibling-FHAD. Our findings highlight the possible effects of sex-specific information bias, participation bias, and confounding bias in FHAD assessment. This has major implications for AD research. Further studies will be required to quantify these effects and propose feasible solutions to minimize and eliminate the biases.

## ACKNOWLEDGMENTS

We sincerely thank all the participants enrolled in the UK Biobank, as well as the investigators contributing to this initiative. We gratefully acknowledge All of Us participants for their contributions, without whom this research would not have been possible. We also thank the National Institutes of Health’s All of Us Research Program for making available the participant data examined in this study.

## DATA AVAILABILITY

The UKB individual data could be obtained by applying on the website (http://www.ukbiobank.ac.uk/). The UKB GWAS data could be downloaded from the Neale Lab (http://www.nealelab.is/uk-biobank) and Oxford Brain Imaging Genetics Server (https://open.win.ox.ac.uk/ukbiobank/big40/). The AoU data in this article were accessed from the All of Us Curated Data Repository version 7 (https://www.researchallofus.org). PRS was calculated using PRScs (https://github.com/getian107/PRScs) and PLINK 1.9 (https://www.cog-genomics.org/plink/1.9/). One-sample MR analyses were performed using OneSampleMR (https://github.com/remlapmot/OneSampleMR) and ivreg (https://github.com/zeileis/ivreg/).

## AUTHOR CONTRIBUTIONS

RP and JH designed the study. EF and JH extracted and preprocessed the data. BCM and GAP provided statistical methodological support. JH conducted the formal analysis and wrote the manuscript. BCM, EF, APM, CHvD, and GAP contributed to the data interpretation. All the authors participated in the critical revision of the manuscript for important intellectual content. RP obtained the primary funding and supervised the study. All authors read and approved the final manuscript.

## CONSENT STATEMENT

The UK Biobank has approval from the North-West multi-centre research ethics committee as a Research Tissue Bank approval. The All of Us Research Program was approved by the All of Us Institutional Review Board. All participants in the two studies provided informed consent at recruitment. The data used in this study were obtained through an application reference No.

58146 for the UK Biobank and through an approved data use agreement for the All of Us Research Program.

