## Supplementary Figures for "Sex differences in the associations of socioeconomic factors and cognitive performance with family history of Alzheimer’s disease"

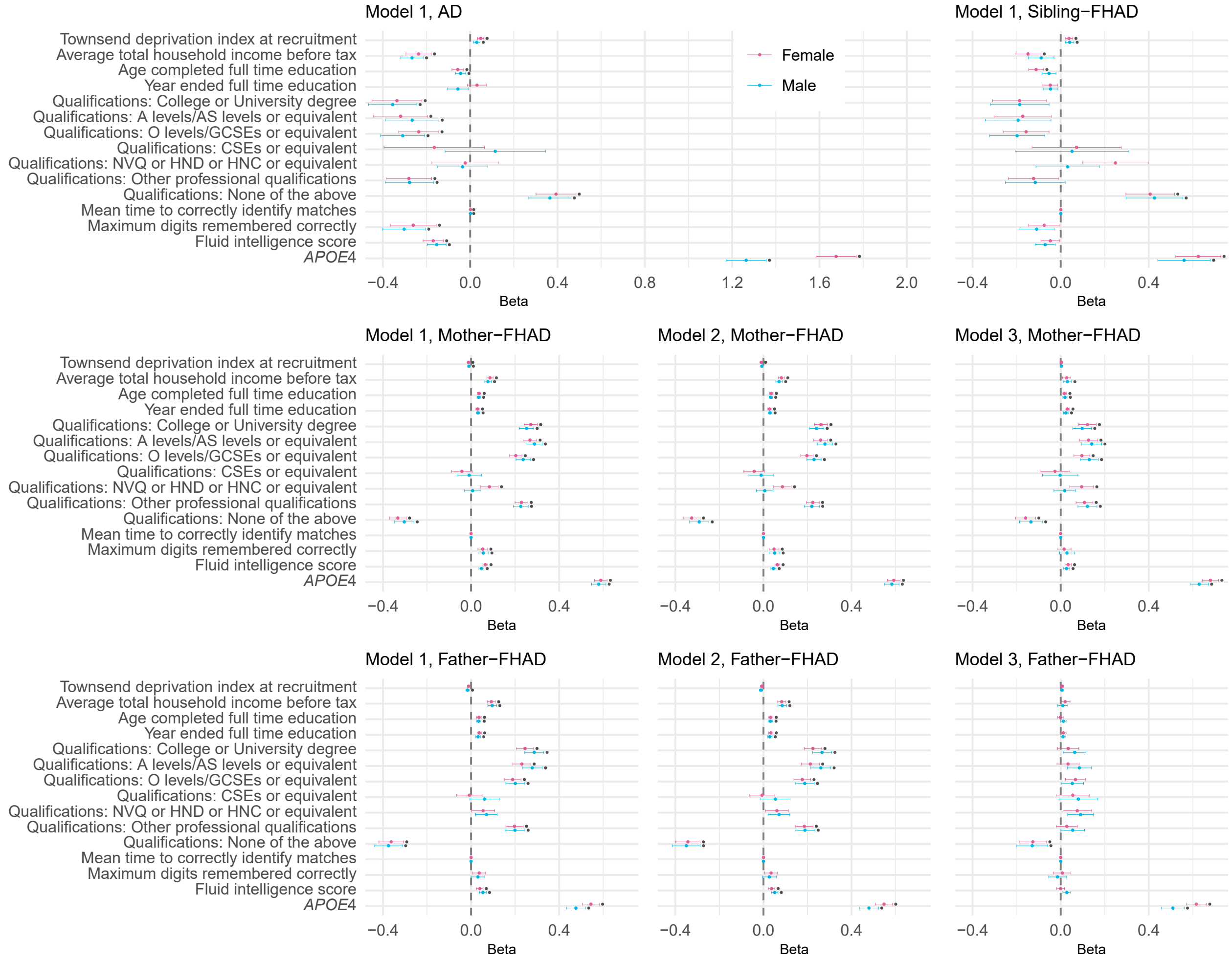


**FIGURE S1 Associations of socioeconomic factors, cognitive performance, and *APOE*4 status with AD and FHAD in the UK Biobank in three models.** In model 1, age and genetically inferred ancestry were included as covariates. In model 2, parental death (father's death for father-FHAD and mother's death for mother-FHAD) was added to covariates already included in model 1. In model 3, the analysis was restricted to participants whose parent died (participants whose father died in father-FHAD analysis and participants whose mother died in mother-FHAD analysis) and included age, genetically inferred ancestry, and parental age at death (father's age at death for father-FHAD and mother's age at death for mother-FHAD). Star symbols indicate associations surviving Bonferroni multiple testing correction. AD: Alzheimer's disease; FHAD: family history of AD.

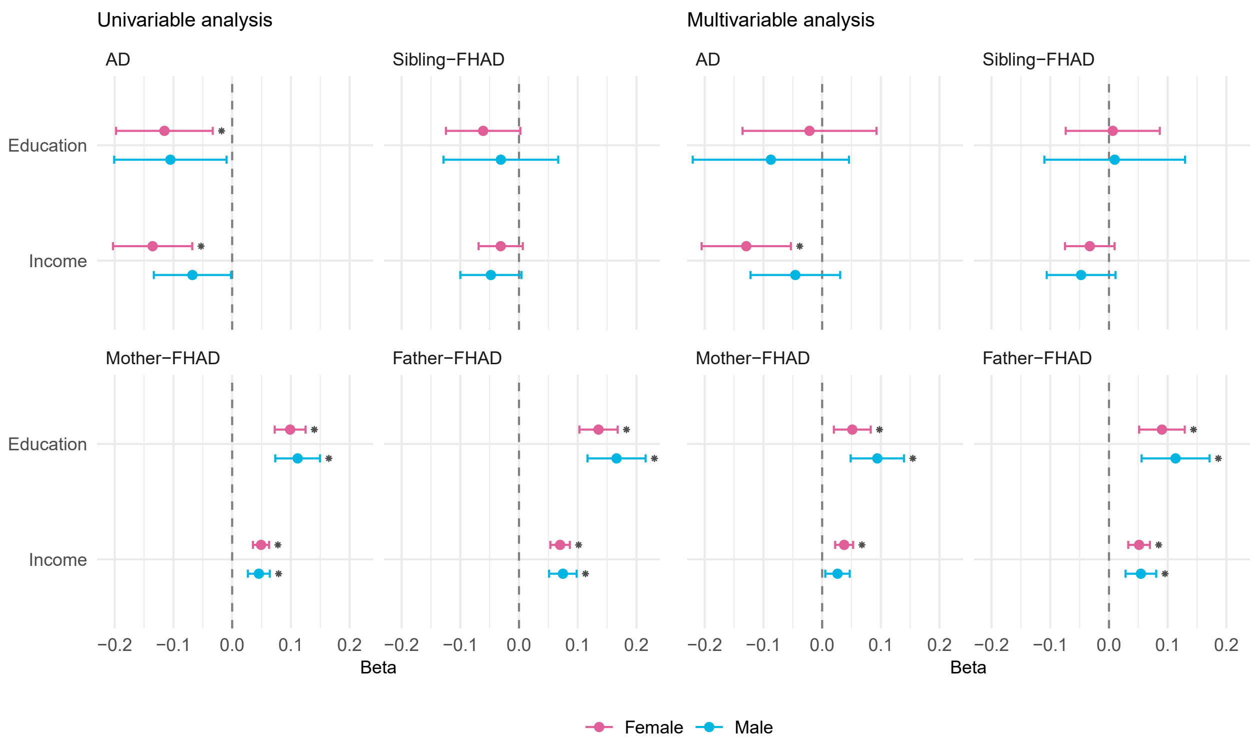


**FIGURE S2 Associations of socioeconomic factors with AD and FHAD in the All of Us Research Program.** Age and genetically inferred ancestry were included as covariates in both univariable and multivariable analyses. Star symbols indicate associations surviving Bonferroni multiple testing correction. AD: Alzheimer's disease; FHAD: family history of AD.
